# Augmenting electronic health record data with social and environmental determinant of health measures to understand regional factors associated with asthma exacerbations

**DOI:** 10.1101/2024.10.24.24316063

**Authors:** Alana Schreibman, Kimberly Lactaoen, Jaehyun Joo, Patrick K. Gleeson, Gary E. Weissman, Andrea J. Apter, Rebecca A. Hubbard, Blanca E. Himes

## Abstract

Electronic health records (EHRs) provide rich data for diverse populations but often lack information on social and environmental determinants of health (SEDH) that are important for the study of complex conditions such as asthma, a chronic inflammatory lung disease. We integrated EHR data with seven SEDH datasets to conduct a retrospective cohort study of 6,656 adults with asthma. Using Penn Medicine encounter data from January 1, 2017 to December 31, 2020, we identified individual-level and spatially-varying factors associated with asthma exacerbations. Black race and prescription of an inhaled corticosteroid were strong risk factors for asthma exacerbations according to a logistic regression model of individual-level risk. A spatial generalized additive model (GAM) identified a hotspot of increased exacerbation risk (mean OR = 1.41, SD 0.14, p < 0.001), and inclusion of EHR-derived variables in the model attenuated the spatial variance in exacerbation odds by 34.0%, while additionally adjusting for the SEDH variables attenuated the spatial variance in exacerbation odds by 66.9%. Additional spatial GAMs adjusted one variable at a time revealed that neighborhood deprivation (OR = 1.05, 95% CI: 1.03, 1.07), Black race (OR = 1.66, 95% CI: 1.44, 1.91), and Medicaid health insurance (OR = 1.30, 95% CI: 1.15, 1.46) contributed most to the spatial variation in exacerbation odds. In spatial GAMs stratified by race, adjusting for neighborhood deprivation and health insurance type did not change the spatial distribution of exacerbation odds. Thus, while some EHR-derived and SEDH variables explained a large proportion of the spatial variance in asthma exacerbations across Philadelphia, a more detailed understanding of SEDH variables that vary by race is necessary to address asthma disparities. More broadly, our findings demonstrate how integration of information on SEDH with EHR data can improve understanding of the combination of risk factors that contribute to complex diseases.

**Author summary:** Electronic health records constitute an important source of data for understanding the health of large and diverse real-world populations, however, they do not routinely capture socioeconomic and environmental factors known to affect health outcomes. We show how electronic health record data can be augmented to include individual measures of air pollution exposures, neighborhood socioeconomic status, and the natural and built environment using patients’ residential addresses to study asthma exacerbations, episodes of worsening disease that remain a major public health challenge in the United States. We found that on an individual patient-level, Black race and prescription of an inhaled corticosteroid were the factors most strongly associated with asthma exacerbations. In contrast, neighborhood deprivation, race, and health insurance type accounted for the most spatial variation in exacerbation risk across Philadelphia. Our findings provide insight into factors that contribute to asthma disparities in our region and present a framework for future efforts to expand the scope of electronic health record data.

## Introduction

Electronic health records (EHRs) are a source of rich patient-level data for large and diverse populations that can be used for research due to their widespread availability [1]. However, EHR data often contain incomplete or low-quality measures of social and environmental determinants of health (SEDH), limiting their utility for the study of complex diseases [2]. Recent efforts to address this limitation have included developing methodologies to integrate external data on the physical, built, and social environment via linkage with patient addresses [3–5], with high-resolution geospatial datasets providing the closest estimate of individual exposures [6]. Integrated EHR and SEDH datasets can be used to understand both individual-level outcomes and patterns of risk across a spatial region, thereby providing insights for both precision medicine and precision public health efforts. Because many environmental exposures pose a greater risk to select groups of people, integrating external information on SEDH with EHR data is also helpful to address health disparities according to race, ethnicity, and socioeconomic status.

Asthma, a chronic disease that is characterized by inflammation and reversible narrowing of the airways, affects over 25 million people or approximately 8% of the United States population [7]. Racial and ethnic disparities in its morbidity and mortality are well known, and those living in poverty are also more likely to have asthma [7–11]. The clinical goals of asthma management are to control patients’ symptoms and minimize long-term risk of lung function decline [12]. This includes preventing asthma exacerbations, episodes of worsening disease which require treatment with systemic steroids [13]. However, despite guideline-directed clinical management, asthma exacerbations remain common, contributing to asthma-related morbidity and mortality as well as higher health care costs and utilization [14,15]. Risk factors for asthma exacerbations in adults include female sex [16], obesity [17], current or past smoking [18], comorbid allergic rhinitis or chronic obstructive pulmonary disease (COPD) [17,19], and a history of previous exacerbations [20]. Observational studies have also found independent associations between asthma exacerbations and exposure to particulate matter (PM), gaseous air pollutants, and to mixtures of pollutants such as traffic-related air pollution (TRAP) [21]. Similarly, associations between asthma exacerbations and living in poverty [22], substandard housing conditions, such as the presence of mold and pests [23,24], and neighborhood “greenness” [25,26] have been documented.

Because asthma exacerbations result from complex interactions among various biological, social, and environmental factors that vary across individuals and geographically, creating generalizable models of exacerbations remains an unachieved goal. Further, because in the United States there is a high correlation between minoritized race or ethnicity, poverty, and harmful environmental exposures, disentangling relationships among key variables is difficult [9,27]. Approaches that model many asthma-related variables in specific regions may lead to the identification of actionable strategies to reduce exacerbations locally, and using EHR data to create these models has the advantage of providing health information for the specific catchment region served by a given healthcare system. Few studies have linked EHRs with a diverse set of SEDH variables to study asthma exacerbations using patient-level data [28–30], and to our knowledge, none have used geospatial analysis techniques to understand the contribution of these factors to the spatial distribution of exacerbation risk. Here, we show how EHR data can be extended to include individualized measures of air pollution exposures, socioeconomic status indices, and measures of the natural and built environment to identify local factors associated with asthma exacerbation risk.

## Methods

A retrospective cohort analysis was performed using de-identified EHR data from Penn Medicine, a large health system that serves the greater Philadelphia area, from encounters dated 1/1/2017-12/31/2020. An overview of our study design is shown in Fig 1.

**Figure 1.**
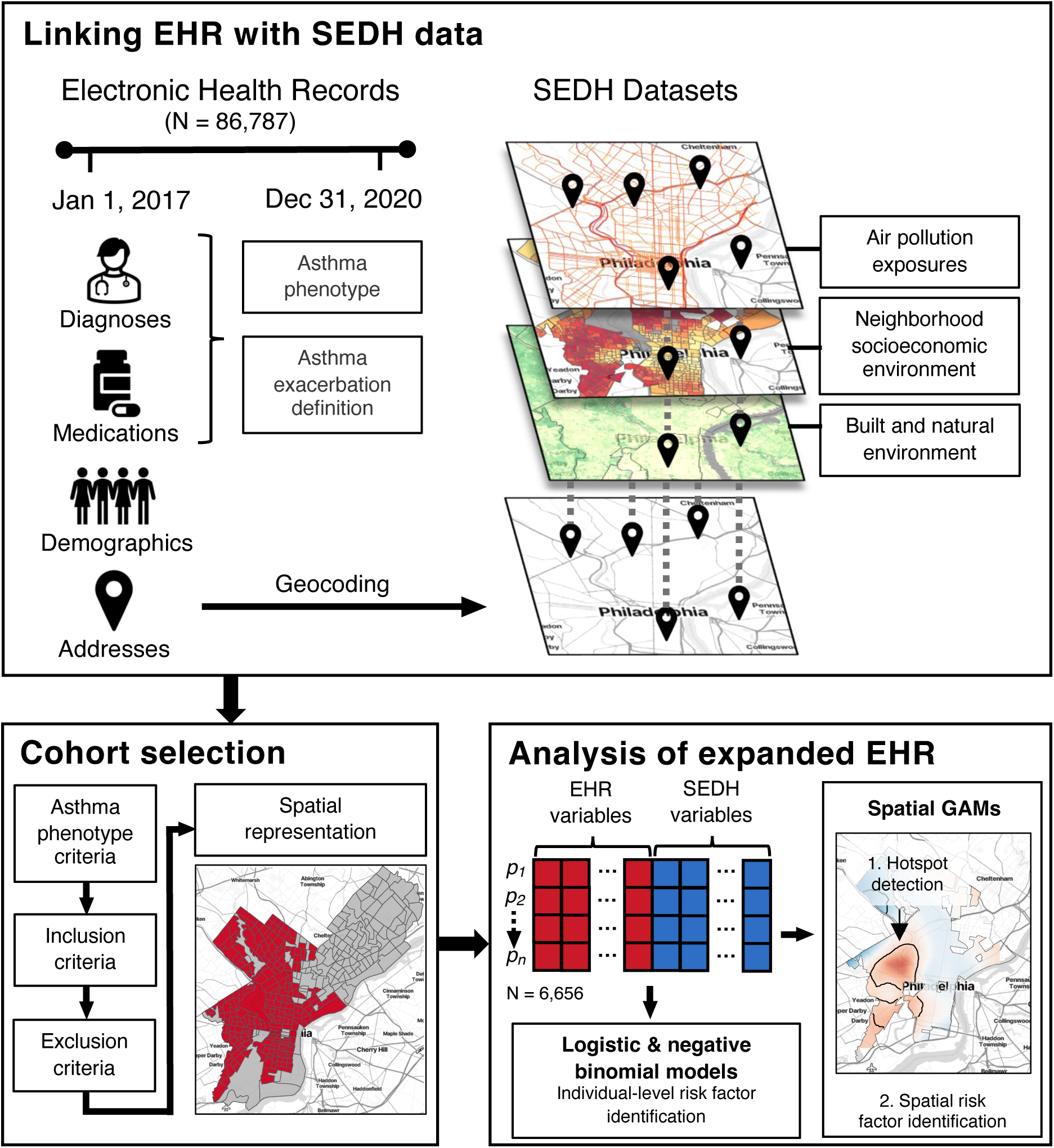
Overview of study design. Graphical overview of study design, including processing and linkage of electronic health record (EHR) and social and environmental determinants of health (SEDH) data, cohort selection including spatial filtering by assessing the representation of the EHR cohort compared to the underlying population, and patient-level and geospatial analyses on the expanded EHR dataset.

### Ethics statement

Our study was approved by the University of Pennsylvania Institutional Review Board (IRB) under protocol number 824789. Formal consent was not obtained, as a waiver of HIPAA Authorization was granted for the conduct of this research.

### Study population

Patient-level encounter data was obtained for adults (i.e. age ≥ 18 years) who had at least one encounter with an International Classification of Diseases (ICD)-10 code for asthma (J45*) and who were prescribed a short-acting β_2_-agonist (SABA) (S1 Table). The most recent residential address for each patient was obtained and geocoded using previously described methods [3]. Demographic, comorbidity, and medication data for encounters during the study period was extracted and used to compute several variables, hereafter referred to as “EHR-derived variables.” These included: age at first encounter, sex, race, ethnicity, body mass index (BMI), health insurance type, smoking status, chronic obstructive pulmonary disease (COPD), allergic rhinitis, a modified Elixhauser score [31], inhaled corticosteroid (ICS) prescription, and years followed (defined as the number of years between first and last encounter). Additional details, including inclusion and exclusion criteria and definitions of the EHR-derived variables, are provided in S1 Text.

### Outcome

Asthma exacerbations were defined as encounters with an oral corticosteroid (OCS) prescription (S1 Table) and either 1) a primary asthma diagnosis code (ICD-10, J45*) for encounters with primary diagnosis codes listed or 2) a nonprimary asthma ICD-10 code for encounters without a primary diagnosis listed but only one or two ICD-10 codes listed. A count of exacerbations during the study period was computed for each patient.

### SEDH data

Seven external datasets were integrated with our EHR dataset via linkage with patient geocodes, creating variables that are hereafter referred to as “SEDH variables.” Additional details on data processing are reported in S1 Text, and dataset sources and spatiotemporal dimensions are summarized in S2 Table. All processed SEDH data is available in <u>S</u>ensor-based <u>A</u>nalysis of <u>P</u>ollution in the <u>Phi</u>ladelphia <u>R</u>egion with Information on <u>N</u>eighborhoods and the <u>E</u>nvironment (SAPPHIRINE), a web application that integrates spatially distributed high-resolution social and environmental data in the greater Philadelphia region to facilitate the conduct of local health studies [32].

#### Air pollution exposures

Average pollutant exposures were assigned to each patient using high-resolution (∼1×1 km^2^) geophysical model estimates. 2017-2019 NO_2_ estimates (in parts per billion by volume, or ppbv) and 2017-2020 PM2.5 estimates (in μg/m^3^) were downloaded from resources reported in Cooper et al. and van Donkelaar et al., respectively, temporally averaged, and linked to the study cohort using bilinear interpolation [33,34]. Exposure to other toxic airborne chemicals from point sources was estimated using the EPA Toxics Release Inventory (TRI) [35]. Total toxic air release exposure by patient was computed as the sum of toxic releases (in kilograms) over the study period within a 1-km circular buffer of each patient’s residential address. Exposure to air pollution from mobile sources was estimated as the sum of the daily vehicle distance traveled (DVDT), a metric computed using traffic data published by the Pennsylvania Department of Transportation, within a 300-m circular buffer [36].

#### Neighborhood socioeconomic environment

Socioeconomic disadvantage for each person was summarized as the Area Deprivation Index (ADI), a validated index computed for each Census block group based on several ACS variables [37]. A higher ADI score indicates greater disadvantage. 2018 ADI data was extracted from the Neighborhood Atlas and was assigned to each patient by block group [38].

#### Built and natural environment

Asthma-related housing code violation data (i.e. pests, water damage, and indoor air contamination) was obtained from a Philadelphia Department of Licenses and Inspections dataset reported by OpenDataPhilly (S3 Table) [39]. For each block group, the number of violations during the study period per 100 people (based on the 2019 ACS population estimates) was computed and assigned to each patient. Vegetation density was summarized as the normalized difference vegetation index (NDVI), an index with values ranging from −1.0 to 1.0 where higher positive values represent higher vegetation density. NDVI was computed in Google Earth Engine using surface reflectance images from the Landsat 8 satellite during the study period and assigned to each patient as the mean value within a 300-m circular buffer [40].

### Statistical analysis

Analyses were conducted in R 4.2 [41].

#### Study area

To minimize bias in our geospatial analyses introduced by uneven spatial density across the Penn Medicine catchment area, we restricted our study region to spatial areas in which the geospatial representativeness of our EHR cohort was adequate compared to the underlying population. Following methods described in Xie et al. [42], we computed a spatial representation ratio (SRR), defined as the ratio between the proportion of our EHR cohort living in each census block group and the proportion of the Philadelphia population living in that block group, as reported by the 2019 American Community Survey (ACS). We defined our study region as contiguous census tracts (i.e. adjacent or separated by a non-residential area such as a park) with a mean SRR of 0.5 or greater. Only patients who resided in this study region were included in analyses.

#### Modeling individual-level risk factors

Chi-squared and Kruskal-Wallis rank sum tests were used in bivariate analyses to assess associations between patient characteristics and asthma exacerbation level (i.e. 0, 1-2, 3-4, 5+) during the study period, and to compare the characteristics of complete cases to individuals with missing data. To identify patient-level factors associated with asthma exacerbations, we fit logistic regression models with asthma exacerbations as a binary case-control (0 vs >0) outcome. This approach was chosen to match the dichotomous outcome used in spatial analyses. Logistic regression models were initially adjusted for EHR-derived variables only, and then adjusted for both EHR-derived and SEDH variables (EHR & SEDH-adjusted). Years followed was included as a covariate in both models to account for variation in the length of available follow-up across patients. Model fit was assessed using the Akaike information criterion (AIC). We checked for multicollinearity by computing Pearson’s correlation coefficients between all EHR and SEDH variables, while selecting White race and Private health insurance type as reference levels for the two nominal categorical variables, and we computed variance inflation factors (VIF) for all independent variables.

#### Sensitivity analysis for individual-level risk factors

We conducted sensitivity analyses of individual-level risk factors by fitting negative binomial regression models with asthma exacerbations represented as a count outcome and comparing results to those of logistic regression models. The negative binomial regression models were adjusted first for EHR-derived variables only and then for EHR & SEDH variables, while including years followed as an offset in each model.

#### Modeling spatial risk factors

To estimate local odds of exacerbation as a function of location, spatial generalized additive models (GAMs) were fit with a binary case-control outcome (0 vs >0 exacerbations) on a grid of points across the study region, using the R *MapGAM* package and previously described methods (S1 Text) [3,43]. Maps of spatial effect predictions were created for the smoothed spatial term of each model, where the spatial odds ratio (OR) at each point represented the ratio between the odds of exacerbation at that point and the median odds across all points. First, a univariable model adjusted only for years followed, hereafter referred to as “unadjusted model”, was fit to identify hotspots and coldspots across the study region. Next, multivariable models adjusted for 1) only EHR-derived variables and 2) EHR & SEDH variables were compared to the unadjusted model by computing the mean and standard deviation (SD) of the spatial ORs at points within any overlapping hotspots, and by computing the percent difference between the variance in ORs across the full study region in the adjusted models and the unadjusted model. To understand the contribution of individual variables to the observed spatial effects, we fit additional models adjusted for each EHR-derived or SEDH variable one variable at a time (all models were also adjusted for years followed) and computed the percent difference between the variance in ORs in these models and the unadjusted model. Model fit was assessed using the AIC.

#### Stratified analysis

We conducted a stratified analysis to further evaluate the association between race and asthma exacerbations. Chi-square and Wilcoxon rank sum tests were used to assess bivariate relationships between race and other variables. To test whether any EHR or SEDH variables that were correlated with race influenced spatial patterns of asthma exacerbation risk independently of race, spatial GAMs adjusted one variable at a time were fit on each race stratum using the same approach described above. Before fitting the models, SRR selection was repeated for each stratum to account for the uneven geographic distribution of race across the initial study region.

## Results

Following selection of patients based on inclusion/exclusion criteria and spatial filtering (S1 Fig), the retrospective study cohort consisted of 6,656 asthma patients, 2,329 of whom had one or more exacerbations (Table 1), with residence in 249 census tracts in Philadelphia (S2 Fig). The spatial distribution of all processed SEDH datasets within the study region is shown in S3 Fig. The EHR-derived variables years followed, age, race, BMI, health insurance type, smoking status, COPD, allergic rhinitis, Elixhauser comorbidity score, and ICS, as well as the SEDH variable ADI were the most significantly associated with exacerbations according to bivariate analyses (p < 0.001) (Table 1). Patients with more exacerbations during the study period were followed for more years, more likely to be aged 35-54, of Black race, or class 2 or class 3 obese, and more likely to have Medicaid health insurance, a history of smoking, COPD, allergic rhinitis, higher Elixhauser comorbidity scores, an ICS prescription, or live in neighborhoods with higher ADI. The proportion of patients in the study cohort who were prescribed controller medications including ICS, leukotriene modifiers, long-acting β_2_-agonists (LABA), and biologic therapies was positively associated with exacerbation count (S4 Table). Bivariate analyses comparing the distribution of characteristics between the study cohort and patients excluded due to missing EHR data found statistically significant differences in years followed, age, sex, health insurance type, COPD, allergic rhinitis, Elixhauser comorbidity score, and ICS (p < 0.001) (S5 Table).

**Table 1.**
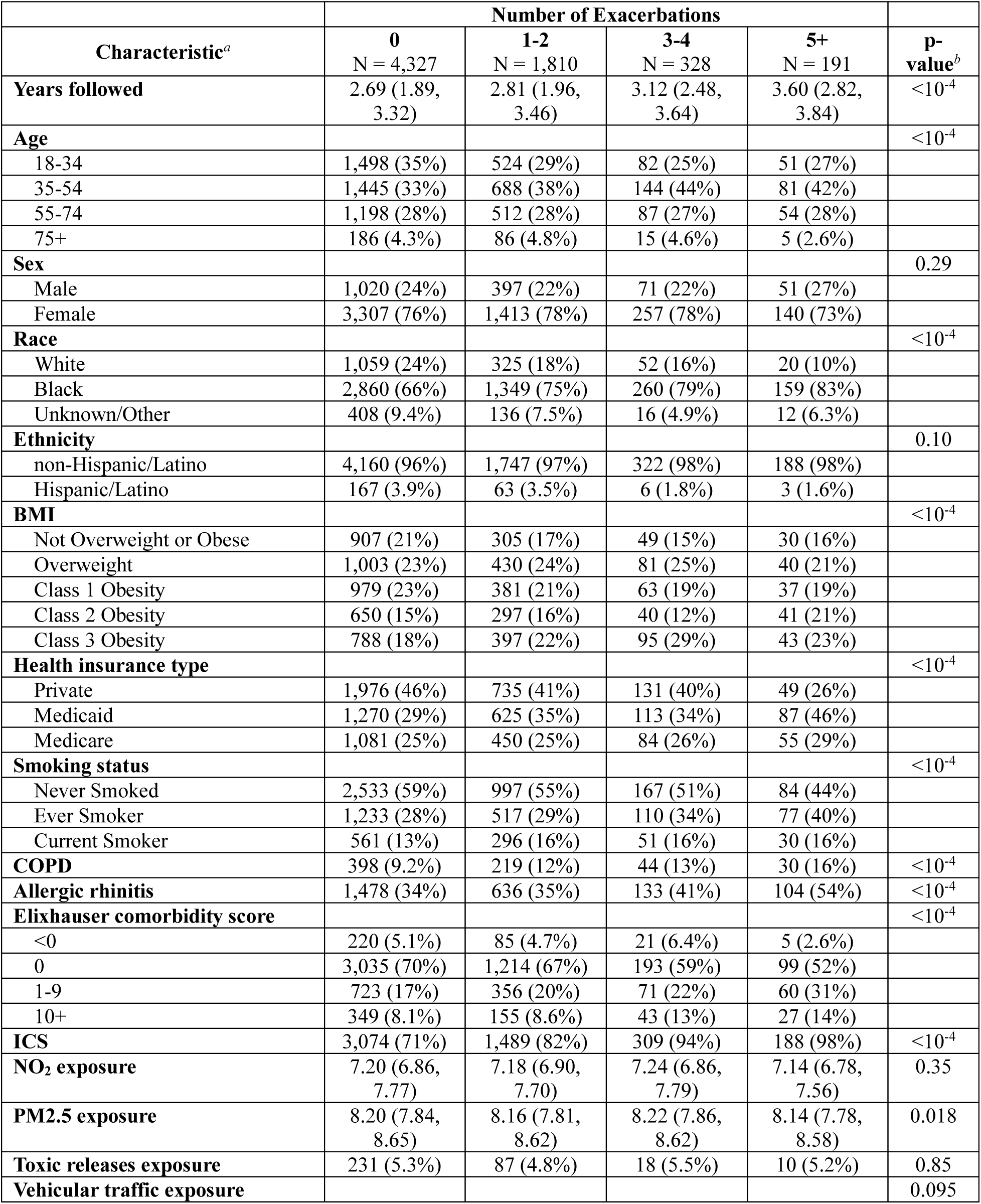
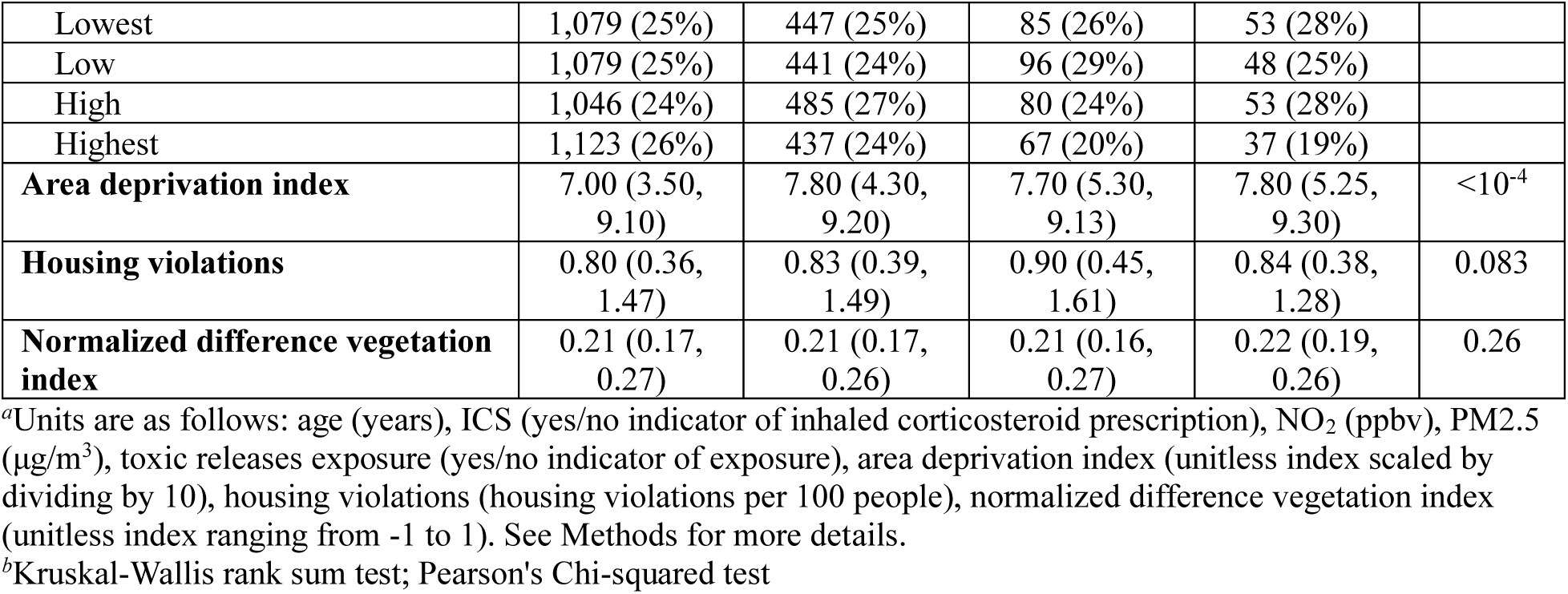
Patient characteristics by exacerbation count levels. Shown are the characteristics of patients according to their number of exacerbations during the study period. For each exacerbation level, the number and percentage of patients are shown for categorical variables, and the Median and Interquartile Range (IQR) are shown for continuous variables.

| Characteristic<sup>a</sup> | Number of Exacerbations |  |  |  | p-value<sup>b</sup> |
| --- | --- | --- | --- | --- | --- |
|  | 0<br>N = 4,327 | 1-2<br>N = 1,810 | 3-4<br>N = 328 | 5+<br>N = 191 |  |
| **Years followed** | 2.69 (1.89, 3.32) | 2.81 (1.96, 3.46) | 3.12 (2.48, 3.64) | 3.60 (2.82, 3.84) | <10<sup>-4</sup> |
| **Age** |  |  |  |  | <10<sup>-4</sup> |
| 18-34 | 1,498 (35%) | 524 (29%) | 82 (25%) | 51 (27%) |  |
| 35-54 | 1,445 (33%) | 688 (38%) | 144 (44%) | 81 (42%) |  |
| 55-74 | 1,198 (28%) | 512 (28%) | 87 (27%) | 54 (28%) |  |
| 75+ | 186 (4.3%) | 86 (4.8%) | 15 (4.6%) | 5 (2.6%) |  |
| **Sex** |  |  |  |  | 0.29 |
| Male | 1,020 (24%) | 397 (22%) | 71 (22%) | 51 (27%) |  |
| Female | 3,307 (76%) | 1,413 (78%) | 257 (78%) | 140 (73%) |  |
| **Race** |  |  |  |  | <10<sup>-4</sup> |
| White | 1,059 (24%) | 325 (18%) | 52 (16%) | 20 (10%) |  |
| Black | 2,860 (66%) | 1,349 (75%) | 260 (79%) | 159 (83%) |  |
| Unknown/Other | 408 (9.4%) | 136 (7.5%) | 16 (4.9%) | 12 (6.3%) |  |
| **Ethnicity** |  |  |  |  | 0.10 |
| non-Hispanic/Latino | 4,160 (96%) | 1,747 (97%) | 322 (98%) | 188 (98%) |  |
| Hispanic/Latino | 167 (3.9%) | 63 (3.5%) | 6 (1.8%) | 3 (1.6%) |  |
| **BMI** |  |  |  |  | <10<sup>-4</sup> |
| Not Overweight or Obese | 907 (21%) | 305 (17%) | 49 (15%) | 30 (16%) |  |
| Overweight | 1,003 (23%) | 430 (24%) | 81 (25%) | 40 (21%) |  |
| Class 1 Obesity | 979 (23%) | 381 (21%) | 63 (19%) | 37 (19%) |  |
| Class 2 Obesity | 650 (15%) | 297 (16%) | 40 (12%) | 41 (21%) |  |
| Class 3 Obesity | 788 (18%) | 397 (22%) | 95 (29%) | 43 (23%) |  |
| **Health insurance type** |  |  |  |  | <10<sup>-4</sup> |
| Private | 1,976 (46%) | 735 (41%) | 131 (40%) | 49 (26%) |  |
| Medicaid | 1,270 (29%) | 625 (35%) | 113 (34%) | 87 (46%) |  |
| Medicare | 1,081 (25%) | 450 (25%) | 84 (26%) | 55 (29%) |  |
| **Smoking status** |  |  |  |  | <10<sup>-4</sup> |
| Never Smoked | 2,533 (59%) | 997 (55%) | 167 (51%) | 84 (44%) |  |
| Ever Smoker | 1,233 (28%) | 517 (29%) | 110 (34%) | 77 (40%) |  |
| Current Smoker | 561 (13%) | 296 (16%) | 51 (16%) | 30 (16%) |  |
| **COPD** | 398 (9.2%) | 219 (12%) | 44 (13%) | 30 (16%) | <10<sup>-4</sup> |
| **Allergic rhinitis** | 1,478 (34%) | 636 (35%) | 133 (41%) | 104 (54%) | <10<sup>-4</sup> |
| **Elixhauser comorbidity score** |  |  |  |  | <10<sup>-4</sup> |
| <0 | 220 (5.1%) | 85 (4.7%) | 21 (6.4%) | 5 (2.6%) |  |
| 0 | 3,035 (70%) | 1,214 (67%) | 193 (59%) | 99 (52%) |  |
| 1-9 | 723 (17%) | 356 (20%) | 71 (22%) | 60 (31%) |  |
| 10+ | 349 (8.1%) | 155 (8.6%) | 43 (13%) | 27 (14%) |  |
| **ICS** | 3,074 (71%) | 1,489 (82%) | 309 (94%) | 188 (98%) | <10<sup>-4</sup> |
| **NO<sub>2</sub> exposure** | 7.20 (6.86, 7.77) | 7.18 (6.90, 7.70) | 7.24 (6.86, 7.79) | 7.14 (6.78, 7.56) | 0.35 |
| **PM<sub>2.5</sub> exposure** | 8.20 (7.84, 8.65) | 8.16 (7.81, 8.62) | 8.22 (7.86, 8.62) | 8.14 (7.78, 8.58) | 0.018 |
| **Toxic releases exposure** | 231 (5.3%) | 87 (4.8%) | 18 (5.5%) | 10 (5.2%) | 0.85 |
| **Vehicular traffic exposure** |  |  |  |  | 0.095 |
| Lowest | 1,079 (25%) | 447 (25%) | 85 (26%) | 53 (28%) |  |
| Low | 1,079 (25%) | 441 (24%) | 96 (29%) | 48 (25%) |  |
| High | 1,046 (24%) | 485 (27%) | 80 (24%) | 53 (28%) |  |
| Highest | 1,123 (26%) | 437 (24%) | 67 (20%) | 37 (19%) |  |
| **Area deprivation index** | 7.00 (3.50, 9.10) | 7.80 (4.30, 9.20) | 7.70 (5.30, 9.13) | 7.80 (5.25, 9.30) | $<10^{-4}$ |
| **Housing violations** | 0.80 (0.36, 1.47) | 0.83 (0.39, 1.49) | 0.90 (0.45, 1.61) | 0.84 (0.38, 1.28) | 0.083 |
| **Normalized difference vegetation index** | 0.21 (0.17, 0.27) | 0.21 (0.17, 0.26) | 0.21 (0.16, 0.27) | 0.22 (0.19, 0.26) | 0.26 |

<sup>a</sup>Units are as follows: age (years), ICS (yes/no indicator of inhaled corticosteroid prescription), NO<sub>2</sub> (ppbv), PM2.5 (µg/m<sup>3</sup>), toxic releases exposure (yes/no indicator of exposure), area deprivation index (unitless index scaled by dividing by 10), housing violations (housing violations per 100 people), normalized difference vegetation index (unitless index ranging from -1 to 1). See Methods for more details.
<sup>b</sup>Kruskal-Wallis rank sum test; Pearson's Chi-squared test

Based on Pearson’s correlation coefficients, there were no strong correlations between any EHR-derived or SEDH variables, except for NO_2_ and PM2.5 (ρ = 0.77) (S4 Fig). Moderate correlations (|ρ| > 0.50) were observed between age and Medicare health insurance, Black race and ADI, and NO_2_ and NDVI. Furthermore, adjusted generalized VIFs for all variables did not exceed 2.00, suggesting that all variables could be included in multivariable models (S6 Table).

### Individual risk factors associated with asthma exacerbations

ORs for the EHR-derived and SEDH variables included in the multivariable logistic regression model are summarized in Table 2. The EHR-derived variables years followed, age 35-54, Black race, and ICS had the strongest positive associations with having at least one exacerbation during the study period (p < 0.001) (Table 2). These effects persisted after additionally adjusting for SEDH variables, and of the SEDH variables in the logistic regression model, only NO_2_ exposure was positively associated with exacerbations (p = 0.0059). Inclusion of the SEDH variables did not improve model fit as determined by AIC (8,320 for both the EHR-adjusted and EHR & SEDH-adjusted models). Sensitivity analyses showed that the risk factors identified by logistic and negative binomial regression models were mostly consistent, with Black race and ICS prescription having the strongest effects between both models (p < 10^−4^), and variables such as Medicaid health insurance and Elixhauser comorbidity score of 1-9 having statistically significant p-values in both models though smaller in the negative binomial regression (p < 0.001) than the logistic regression (p < 0.05) (S7 Table). However, the negative binomial regression model did not identify statistically significant associations between asthma exacerbations and age 35-54 or NO_2_ exposure as did the logistic regression, although the directions of effect were consistent in both models. In addition, the negative binomial regression model identified an association with the SEDH variable housing code violations (p = 0.045) that was not present in the logistic regression model.

**Table 2.** Individual-level asthma exacerbation risk factors in multivariable logistic regression models. Shown are the adjusted odds ratios (ORs), 95% confidence intervals (CIs), and p-values for logistic regression models of asthma exacerbations as a dichotomous outcome adjusted for EHR-derived variables only and for both EHR and SEDH variables.

| Characteristic<sup>a</sup> | EHR-adjusted |  |  | EHR & SEDH-adjusted |  |  |
| --- | --- | --- | --- | --- | --- | --- |
|  | OR | 95% CI | p-value | OR | 95% CI | p-value |
| **Years followed** | 1.16 | 1.09, 1.24 | <10<sup>-4</sup> | 1.17 | 1.09, 1.24 | <10<sup>-4</sup> |
| **Age** |  |  |  |  |  |  |
| 18-34 | — | — |  | — | — |  |
| 35-54 | 1.26 | 1.11, 1.44 | 5.2x10<sup>-4</sup> | 1.27 | 1.12, 1.45 | 3.6x10<sup>-4</sup> |
| 55-74 | 1.08 | 0.92, 1.28 | 0.35 | 1.09 | 0.92, 1.29 | 0.30 |
| 75+ | 1.29 | 0.96, 1.74 | 0.090 | 1.33 | 0.99, 1.79 | 0.060 |
| **Sex** |  |  |  |  |  |  |
| Male | — | — |  | — | — |  |
| Female | 0.99 | 0.87, 1.12 | 0.82 | 0.98 | 0.87, 1.12 | 0.79 |
| **Race** |  |  |  |  |  |  |
| White | — | — |  | — | — |  |
| Black | 1.49 | 1.29, 1.72 | <10<sup>-4</sup> | 1.52 | 1.28, 1.81 | <10<sup>-4</sup> |
| Unknown/Other | 1.01 | 0.79, 1.28 | 0.95 | 1.01 | 0.79, 1.29 | 0.93 |
| **Ethnicity** |  |  |  |  |  |  |
| Non-Hispanic/Latino | — | — |  | — | — |  |
| Hispanic/Latino | 0.99 | 0.71, 1.36 | 0.95 | 0.98 | 0.71, 1.35 | 0.92 |
| **BMI** |  |  |  |  |  |  |
| Not Overweight or Obese | — | — |  | — | — |  |
| Overweight | 1.17 | 0.99, 1.38 | 0.059 | 1.18 | 1.00, 1.39 | 0.054 |
| Class 1 Obesity | 0.97 | 0.82, 1.15 | 0.71 | 0.98 | 0.82, 1.16 | 0.80 |
| Class 2 Obesity | 1.09 | 0.90, 1.31 | 0.39 | 1.09 | 0.90, 1.31 | 0.39 |
| Class 3 Obesity | 1.24 | 1.04, 1.48 | 0.016 | 1.25 | 1.04, 1.49 | 0.015 |
| **Health insurance type** |  |  |  |  |  |  |
| Private | — | — |  | — | — |  |
| Medicaid | 1.22 | 1.07, 1.39 | 0.0034 | 1.19 | 1.04, 1.36 | 0.012 |
| Medicare | 0.90 | 0.77, 1.06 | 0.21 | 0.89 | 0.76, 1.04 | 0.15 |
| **Smoking status** |  |  |  |  |  |  |
| Never Smoked | — | — |  | — | — |  |
| Ever Smoker | 1.04 | 0.92, 1.18 | 0.51 | 1.04 | 0.92, 1.18 | 0.56 |
| Current Smoker | 1.17 | 0.99, 1.37 | 0.059 | 1.16 | 0.98, 1.36 | 0.077 |
| **COPD** |  |  |  |  |  |  |
| No | — | — |  | — | — |  |
| Yes | 1.14 | 0.95, 1.36 | 0.17 | 1.11 | 0.93, 1.34 | 0.24 |
| **Allergic rhinitis** |  |  |  |  |  |  |
| No | — | — |  | — | — |  |
| Yes | 1.09 | 0.97, 1.22 | 0.13 | 1.09 | 0.98, 1.22 | 0.11 |
| **Elixhauser comorbidity score** |  |  |  |  |  |  |
| <0 | — | — |  | — | — |  |
| 0 | 1.20 | 0.94, 1.54 | 0.14 | 1.20 | 0.94, 1.54 | 0.14 |
| 1-9 | 1.40 | 1.08, 1.83 | 0.012 | 1.39 | 1.07, 1.82 | 0.015 |
| 10+ | 1.20 | 0.90, 1.62 | 0.22 | 1.19 | 0.89, 1.61 | 0.25 |
| **ICS** |  |  |  |  |  |  |
| No | — | — |  | — | — |  |
| Yes | 2.19 | 1.91, 2.51 | <10<sup>-4</sup> | 2.20 | 1.92, 2.52 | <10<sup>-4</sup> |
| **NO<sub>2</sub> exposure** |  |  |  | 1.27 | 1.07, 1.51 | 0.0059 |
| **PM<sub>2.5</sub> exposure** |  |  |  | 0.89 | 0.72, 1.09 | 0.25 |
| **Toxic releases exposure** |  |  |  |  |  |  |
| No |  |  |  | — | — |  |
| Yes |  |  |  | 1.07 | 0.83, 1.37 | 0.60 |
| **Vehicular traffic exposure** |  |  |  |  |  |  |
| Lowest |  |  |  | — | — |  |
| Low |  |  |  | 1.01 | 0.87, 1.17 | 0.86 |
| High |  |  |  | 1.10 | 0.95, 1.28 | 0.19 |
| Highest |  |  |  | 0.95 | 0.82, 1.11 | 0.54 |
| **Area deprivation index** |  |  |  | 1.02 | 0.99, 1.04 | 0.22 |
| **Housing violations** |  |  |  | 0.96 | 0.90, 1.01 | 0.11 |
| **Normalized difference vegetation index** |  |  |  | 1.08 | 0.47, 2.44 | 0.86 |
| **AIC** | 8,320 |  |  | 8,320 |  |  |

<sup>a</sup>Units are as follows: age (years), ICS (yes/no indicator of inhaled corticosteroid prescription), NO<sub>2</sub> (ppbv), PM2.5 (µg/m<sup>3</sup>), toxic releases exposure (yes/no indicator of exposure), area deprivation index (unitless index scaled by dividing by 10), housing violations (housing violations per 100 people), normalized difference vegetation index (unitless index ranging from -1 to 1). See Methods for more details.

### Spatial risk factors associated with exacerbations

Maps of ORs across the study region for the unadjusted, EHR-adjusted, and EHR & SEDH-adjusted spatial GAMs are shown in Fig 2. In the unadjusted model, the global test of the null hypothesis that asthma exacerbations were not associated with geographic location was significant (p < 0.001) (Fig 2A). Local tests identified a statistically significant hotspot of exacerbations (p < 0.01) in West and South Philadelphia with a mean spatial OR of 1.41 (SD 0.14). In the model adjusted for EHR-derived variables (Fig 2B) the global test statistic remained significant (p < 0.001). Local tests again identified a hotspot (p < 0.01) in West and South Philadelphia which had a decreased mean spatial OR of 1.27 (SD 0.055). In this EHR-adjusted model, the variance in spatial ORs across the study region was 34.0% lower than the variance of the unadjusted model (S5 Fig). In the model adjusted for both EHR-derived and SEDH variables, the global test statistic remained significant (p < 0.001, Fig 2C) and local tests identified a hotspot (p < 0.01) in West Philadelphia that overlapped geographically with the one in the other models, although of a smaller area and with a smaller mean spatial OR of 1.24 (SD 0.042) compared to the EHR-adjusted model. The variance in spatial ORs in the EHR & SEDH-adjusted model was strongly attenuated (66.9% lower than that of the unadjusted model), suggesting that these variables partially explained the spatial correlation of exacerbations (S5 Fig). The ORs for the other terms included in the spatial GAMs (i.e., the EHR-derived and SEDH variables) are summarized in Table 3. All variables (i.e., years followed, age 35-54, Black race, class 3 obesity, Medicaid health insurance, Elixhauser score 1-9, ICS) that were significant in the multivariable logistic regression models (Table 2) were also significant in the spatial GAMs (Table 3), except for NO_2_ exposure, which was significant in the logistic regression model but not in the spatial GAM.

**Figure 2.**
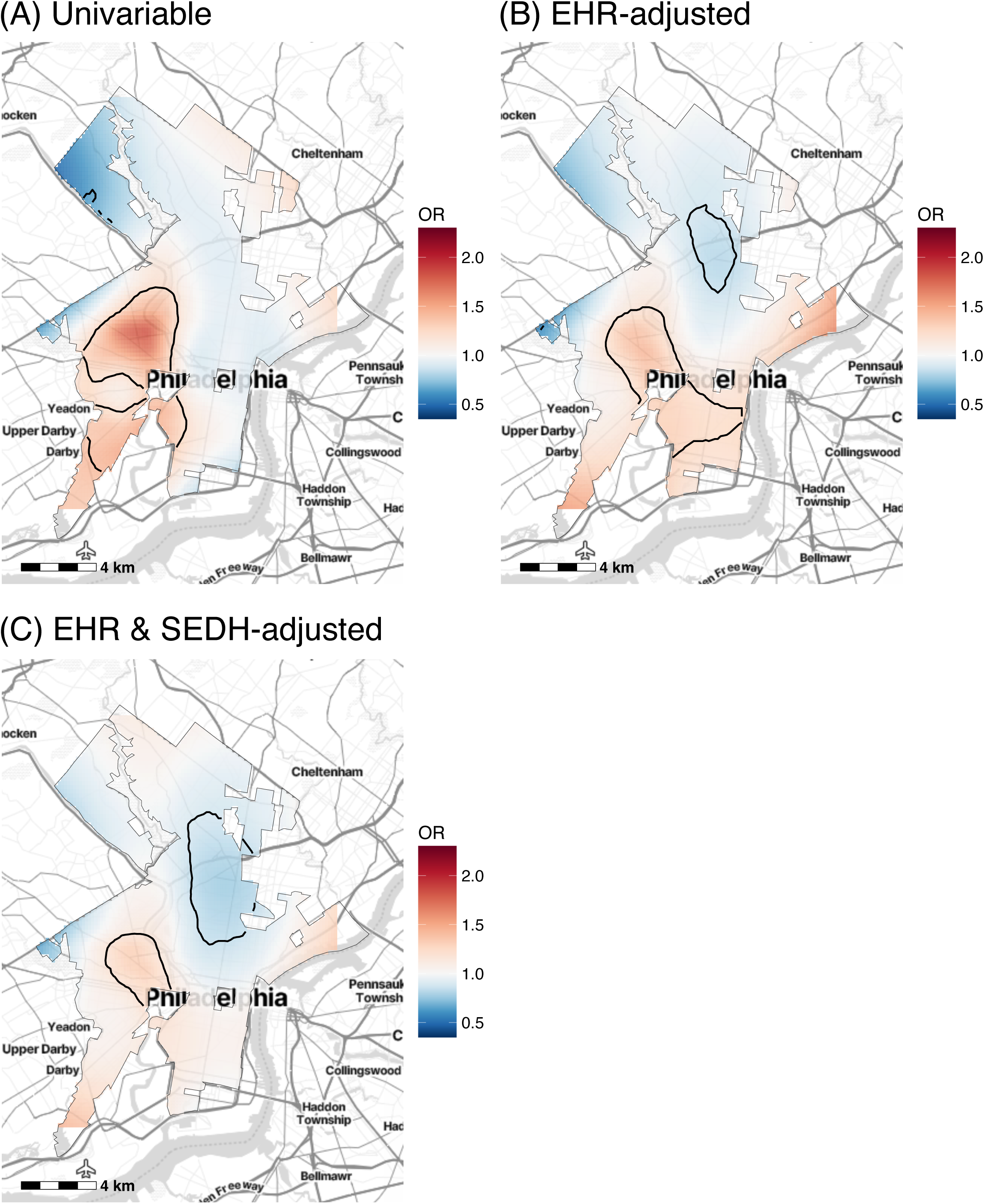
Spatial odds ratios (ORs) of exacerbations before and after adjusting for EHR-derived and SEDH variables. (A) Unadjusted spatial GAM (adjusted only for years followed). (B) Spatial GAM adjusted for EHR-derived variables only. (C) Spatial GAM adjusted for both EHR-derived and SEDH variables. Base maps were created using the Stamen Design from Stadia Maps.

**Table 3.** Spatial asthma exacerbation risk factors in multivariable spatial GAMs. Shown are the adjusted odds ratios (ORs), 95% confidence intervals (CIs), and p-values for spatial GAMs of asthma exacerbations as a dichotomous outcome adjusted for EHR-derived variables only and for both EHR-derived and SEDH variables.

|  | EHR-adjusted |  |  | EHR & SEDH-adjusted |  |  |
| --- | --- | --- | --- | --- | --- | --- |
| Characteristic<sup>a</sup> | OR | 95% CI | p-value | OR | 95% CI | p-value |
| **Years followed** | 1.17 | 1.10, 1.25 | <10<sup>-4</sup> | 1.17 | 1.09, 1.25 | <10<sup>-4</sup> |
| **Age** |  |  |  |  |  |  |
| 18-34 | — | — | — | — | — | — |
| 35-54 | 1.28 | 1.12, 1.46 | 3.3x10<sup>-4</sup> | 1.27 | 1.12, 1.46 | 3.5x10<sup>-4</sup> |
| 55-74 | 1.08 | 0.91, 1.27 | 0.39 | 1.08 | 0.91, 1.27 | 0.38 |
| 75+ | 1.31 | 0.97, 1.76 | 0.078 | 1.32 | 0.98, 1.78 | 0.070 |
| **Sex** |  |  |  |  |  |  |
| Male | — | — | — | — | — | — |
| Female | 0.99 | 0.87, 1.12 | 0.85 | 0.99 | 0.87, 1.12 | 0.84 |
| **Race** |  |  |  |  |  |  |
| White | — | — | — | — | — | — |
| Black | 1.58 | 1.35, 1.84 | <10<sup>-4</sup> | 1.55 | 1.30, 1.85 | <10<sup>-4</sup> |
| Unknown/Other | 1.03 | 0.81, 1.31 | 0.81 | 1.02 | 0.79, 1.30 | 0.90 |
| **Ethnicity** |  |  |  |  |  |  |
| non-Hispanic/Latino | — | — | — | — | — | — |
| Hispanic/Latino | 1.03 | 0.74, 1.42 | 0.87 | 1.02 | 0.74, 1.41 | 0.91 |
| **BMI** |  |  |  |  |  |  |
| Not Overweight or Obese | — | — | — | — | — | — |
| Overweight | 1.18 | 1.01, 1.40 | 0.043 | 1.18 | 1.00, 1.39 | 0.047 |
| Class 1 Obesity | 0.98 | 0.83, 1.16 | 0.82 | 0.98 | 0.83, 1.16 | 0.81 |
| Class 2 Obesity | 1.09 | 0.90, 1.31 | 0.37 | 1.09 | 0.90, 1.31 | 0.39 |
| Class 3 Obesity | 1.26 | 1.05, 1.50 | 0.011 | 1.25 | 1.05, 1.50 | 0.013 |
| **Health insurance type** |  |  |  |  |  |  |
| Private | — | — | — | — | — | — |
| Medicaid | 1.18 | 1.03, 1.35 | 0.014 | 1.18 | 1.03, 1.35 | 0.018 |
| Medicare | 0.89 | 0.76, 1.05 | 0.16 | 0.89 | 0.76, 1.04 | 0.15 |
| **Smoking status** |  |  |  |  |  |  |
| Never Smoked | — | — | — | — | — | — |
| Ever Smoker | 1.04 | 0.91, 1.17 | 0.57 | 1.03 | 0.91, 1.17 | 0.59 |
| Current Smoker | 1.15 | 0.98, 1.35 | 0.094 | 1.15 | 0.98, 1.35 | 0.095 |
| **COPD** |  |  |  |  |  |  |
| No | — | — | — | — | — | — |
| Yes | 1.10 | 0.92, 1.32 | 0.30 | 1.10 | 0.92, 1.32 | 0.29 |
| **Allergic rhinitis** |  |  |  |  |  |  |
| No | — | — | — | — | — | — |
| Yes | 1.09 | 0.98, 1.22 | 0.12 | 1.10 | 0.98, 1.22 | 0.11 |
| **Elixhauser comorbidity score** |  |  |  |  |  |  |
| <0 | — | — | — | — | — | — |
| 0 | 1.20 | 0.94, 1.54 | 0.14 | 1.20 | 0.94, 1.53 | 0.15 |
| 1-9 | 1.39 | 1.06, 1.81 | 0.016 | 1.38 | 1.06, 1.80 | 0.018 |
| 10+ | 1.18 | 0.87, 1.59 | 0.28 | 1.17 | 0.87, 1.58 | 0.29 |
| **ICS** |  |  |  |  |  |  |
| No | — | — | — | — | — | — |
| Yes | 2.22 | 1.93, 2.54 | <10<sup>-4</sup> | 2.21 | 1.93, 2.54 | <10<sup>-4</sup> |
| **NO<sub>2</sub> exposure** |  |  |  | 1.08 | 0.82, 1.43 | 0.58 |
| **PM<sub>2.5</sub> exposure** |  |  |  | 1.08 | 0.81, 1.45 | 0.59 |
| **Toxic releases exposure** |  |  |  |  |  |  |
| No |  |  |  | — | — | — |
| Yes |  |  |  | 1.06 | 0.82, 1.37 | 0.65 |
| **Vehicular traffic exposure** |  |  |  |  |  |  |
| Lowest |  |  |  | — | — | — |
| Low |  |  |  | 1.02 | 0.88, 1.18 | 0.76 |
| High |  |  |  | 1.10 | 0.95, 1.27 | 0.22 |
| Highest |  |  |  | 0.97 | 0.83, 1.13 | 0.66 |
| **Area deprivation index** |  |  |  | 1.01 | 0.99, 1.04 | 0.32 |
| **Housing violations** |  |  |  | 0.97 | 0.91, 1.02 | 0.23 |
| **Normalized difference vegetation index** |  |  |  | 0.99 | 0.43, 2.27 | 0.97 |
| **AIC<sup>b</sup>** | 8,293 |  |  | 8,307 |  |  |

<sup>a</sup>Units are as follows: age (years), ICS (yes/no indicator of inhaled corticosteroid prescription), NO<sub>2</sub> (ppbv), PM2.5 (µg/m<sup>3</sup>), toxic releases exposure (yes/no indicator of exposure), area deprivation index (unitless index scaled by dividing by 10), housing violations (housing violations per 100 people), normalized difference vegetation index (unitless index ranging from -1 to 1). See Methods for more details.
<sup>b</sup>The AIC of the unadjusted model was 8,576.

In the spatial GAM models adjusted one variable at a time, ADI, race, and health insurance type most strongly attenuated the variation in spatial ORs (Fig 3), individually accounting for 55.2%, 38.5%, and 26.5%, respectively, of the variation in the unadjusted model (S5 Fig). In these models, each variable was positively associated with exacerbations (p < 0.001): ADI with OR = 1.05 (95% CI: 1.03, 1.07); Black race with OR = 1.66 (95% CI: 1.44, 1.91); and Medicaid health insurance with OR = 1.30 (95% CI: 1.15, 1.46) (S8 Table). The spatial distribution of these variables followed similar patterns: high ADI and high density of Black patients and patients with Medicaid insurance cooccurred in West, North, and South Philadelphia (Fig 3). No other EHR (S6 Fig) or SEDH (S7 Fig) variable attenuated the hotspot area or its effect size, however, adjustment for NO_2_ levels resulted in an expansion of a coldspot area (S7A Fig).

**Figure 3.**
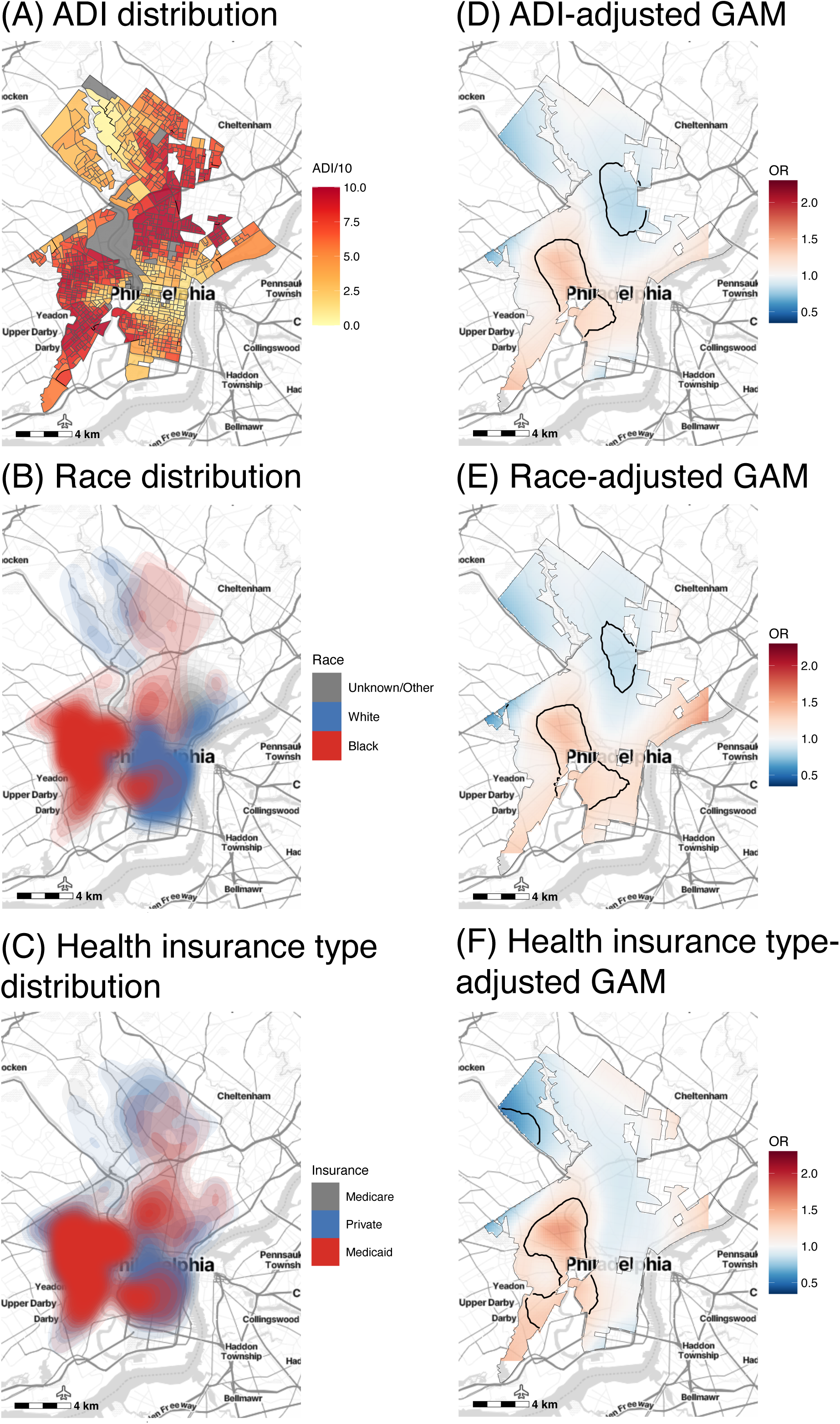
Spatial distribution of individual variables that most strongly attenuated the spatial odds ratios (ORs) of exacerbations along with corresponding spatial GAM results adjusted for these individual variables. Spatial distribution in the study region of (A) the area deprivation index (ADI), (B) race, and (C) health insurance type of patients. Corresponding spatial GAMs adjusted only for years followed and (D) ADI, (E) race, or (F) health insurance type.

### Stratified analysis by race

Bivariate analysis comparing the distribution of all EHR and SEDH characteristics between patients of Black and White race found statistically significant differences for all variables except ethnicity and ICS (Table 4). Given that ADI, race, and health insurance type had the strongest relationship with asthma exacerbations in spatial analyses, and their spatial distributions were similar, we performed stratified spatial analyses by race to determine whether ADI and health insurance status remained significantly associated with asthma exacerbations within race-stratified groups. After applying SRR inclusion criteria in strata according to race, an additional 265 Black patients were excluded from the stratified spatial analysis, resulting in a sample size of 4,363 patients (Fig 4A). The unadjusted spatial GAM for patients of Black race had a significant global test statistic (p < 0.001) and local tests identified hotspots in West and South Philadelphia consistent with results of the full cohort (Fig 4B). In contrast to spatial analyses in the full cohort, adjusting for ADI and health insurance separately for Black patients did not attenuate the spatial variance in ORs, instead increasing the variance by 4.70% and 2.79%, respectively, compared to the unadjusted model (Fig 4C and 4D). In patients of White race, 73 additional patients were removed after applying SRR inclusion criteria, resulting in 1,383 patients included in the spatial model for which the global test statistic was no longer significant (p = 0.098, S8 Fig).

**Table 4.**
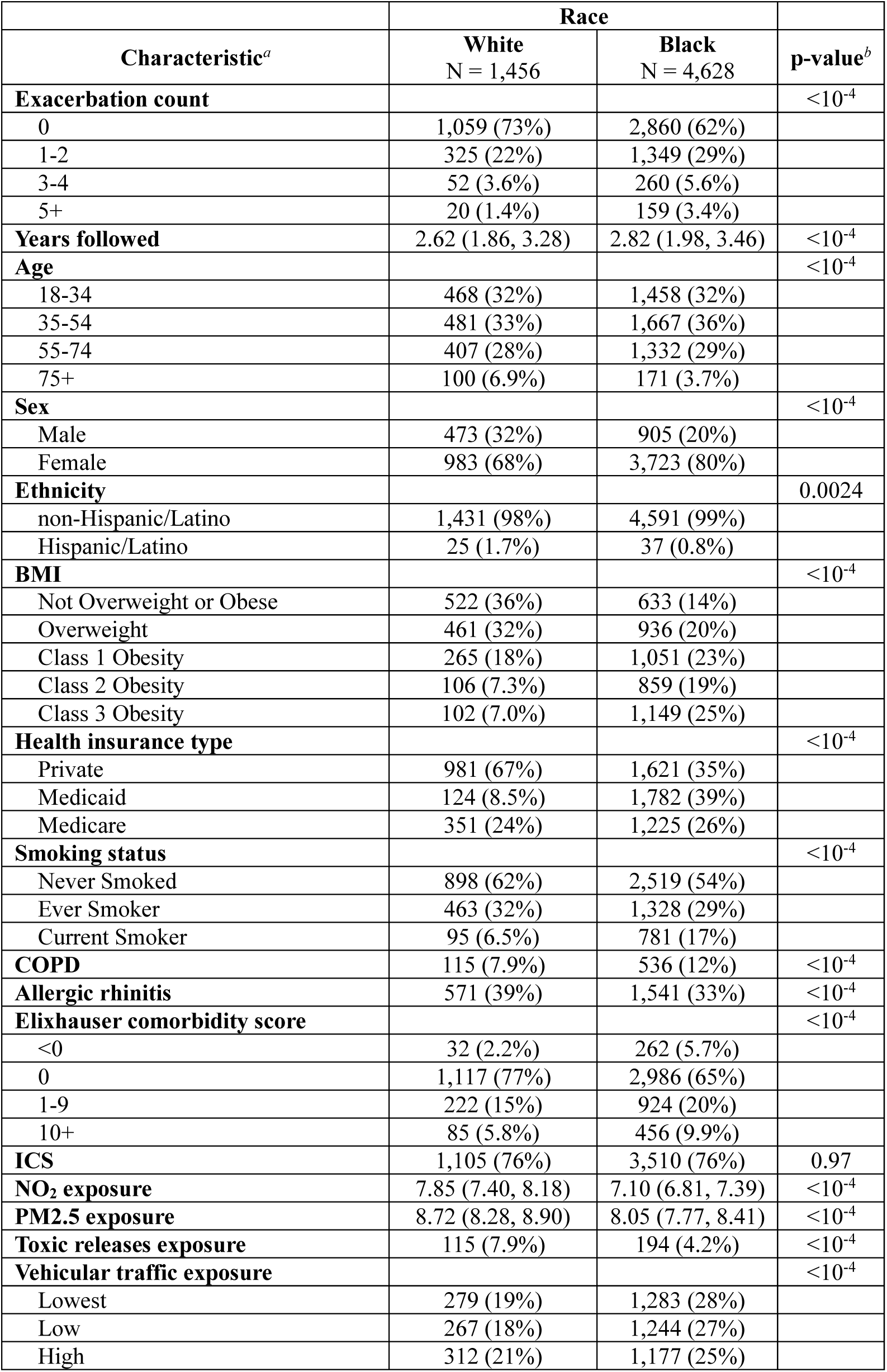
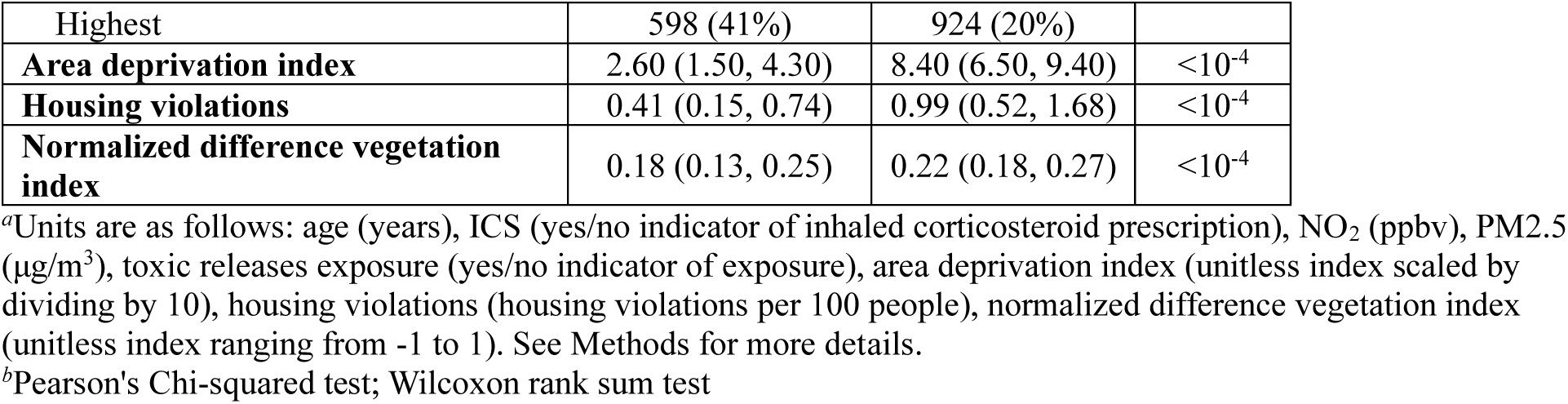
Patient characteristics by race. Shown are the number and percentage of patients in each level for categorical variables, and the Median and Interquartile Range (IQR) for continuous variables in patients of White race versus Black race.

**Figure 4.**
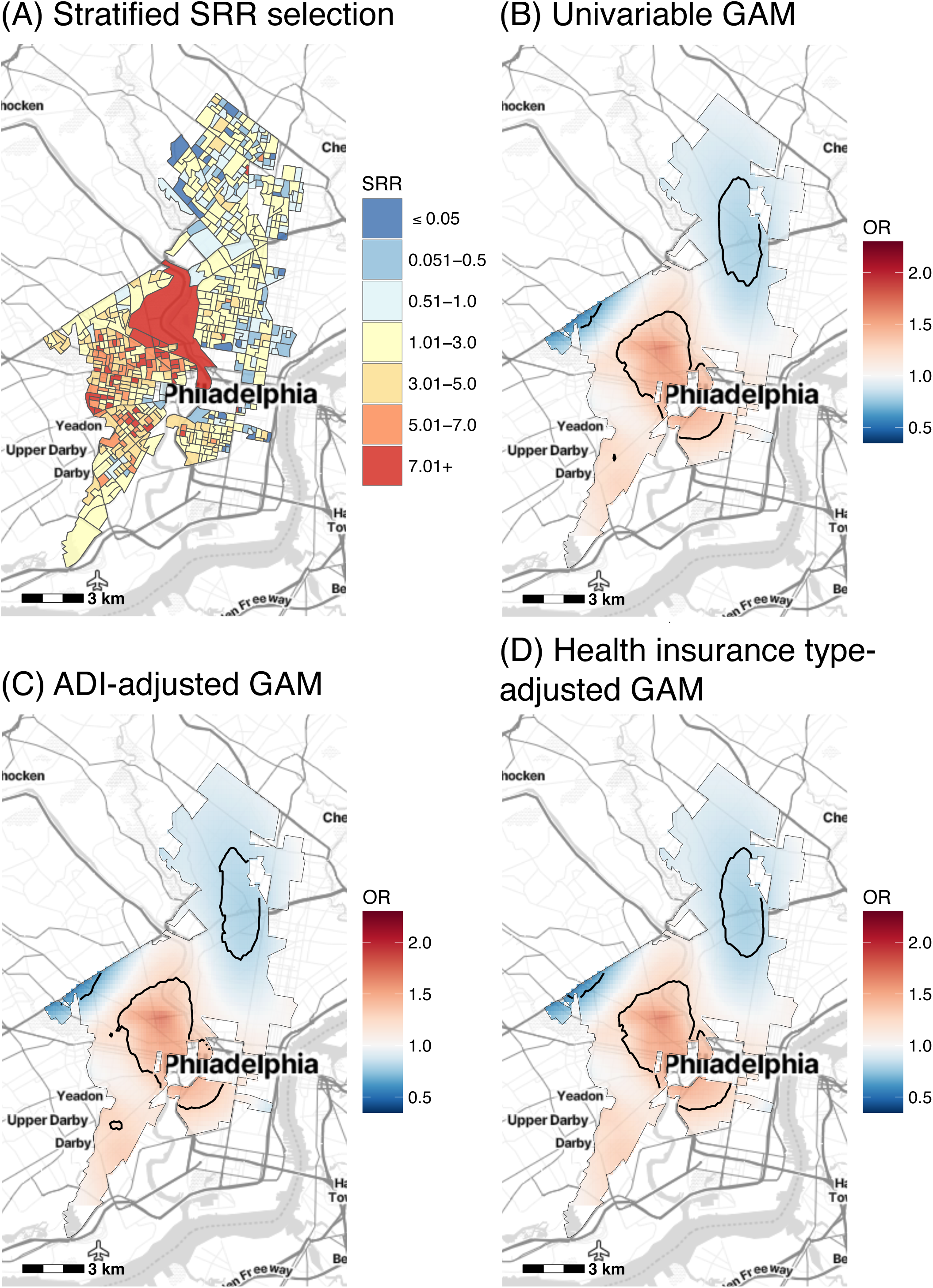
Spatial odds ratios (ORs) of exacerbations among Black patients along with the effects of ADI and health insurance type on this distribution. (A) SRR values for the updated study region used in spatial GAMs for patients of Black race only (SRR = 1 indicates no representativeness bias). (B) Unadjusted spatial GAM (adjusted only for years followed) for patients of Black race (N = 4,363). Spatial GAMs adjusted additionally for (C) area deprivation index (ADI) and (D) health insurance type. Base maps were created using the Stamen Design from Stadia Maps.

## Discussion

Our analysis of individual-level risk factors found that Black race and ICS prescription had the strongest positive associations with asthma exacerbations, as determined by individual-level logistic regression and spatial GAM models. In our cohort of Penn Medicine asthma patients, 66% with no exacerbations were Black compared to 83% with 5+ exacerbations, consistent with known racial disparities in asthma and observations in past Penn Medicine cohorts [44,45]. With regard to the observed association between exacerbations and ICS prescription, international asthma management guidelines underwent a major shift in 2019, recommending ICS as part of the first-line treatment for all asthma patients [46]. This shift was not reflected in our cohort, with only 76.0% of patients prescribed ICS. We observed exacerbations in patients in our cohort regardless of whether they had an ICS prescription, however, the strong association between ICS and exacerbations suggests that patients with more severe asthma were prescribed ICS more frequently than those with milder asthma. Our sensitivity analysis found that logistic regression identified positive associations between age 35-54 and NO_2_ exposure that were not observed in a negative binomial regression model, suggesting that these factors were associated with risk of having at least one exacerbation compared to none, but not with having a higher count of exacerbations.

Our spatial analysis of patient-level data revealed several important insights. First, we observed that asthma exacerbation risk across Philadelphia was spatially correlated and identified a hotspot with 41% higher odds of exacerbation compared to the median across the study region. This finding is consistent with the results of community-based pediatric asthma screening in Philadelphia, which have found that local asthma prevalence can vary significantly from regional or national estimates [47]. Our findings are also consistent with studies that have assessed spatial heterogeneity of pediatric and adult asthma data in other United States metropolitan areas, albeit with less granular spatial resolution. Zárate et al. observed statistically significant spatial patterning of asthma-related emergency department visits across census tracts in Central Texas [48], Grunwell et al. identified a group of contiguous census tracts in the State of Georgia with high rates of admission to the pediatric intensive care unit for asthma [49], and Harris et al. and Corburn et al. identified a statistically significant cluster of zip codes in St. Louis, Missouri and of census tracts in New York City, respectively, with elevated pediatric asthma hospitalization rates [50,51]. Our observation of a West and South Philadelphia hotspot of exacerbation risk is also consistent with analyses of past Penn Medicine cohorts [3,4] that focused on validating methods for augmenting EHR datasets rather than identifying factors associated with the hotspots.

The factors we found to be associated with asthma exacerbations according to EHR- and EHR & SEDH-adjusted spatial GAM models were largely consistent with individual-level logistic regression findings, but the spatial analysis provided an improved context to understand the individual-level results. Adjusting for all EHR and SEDH variables decreased the variance in spatial ORs by 66.9%, indicating that these variables together accounted for a large proportion of the spatial variance in exacerbation odds. By adjusting the spatial model one variable at a time, we found that ADI, race, and health insurance type most attenuated the hotspot area and effect size by reducing the variance of spatial ORs, suggesting that these variables were the most influential in determining the spatial distribution of exacerbation risk. Our findings are consistent with known asthma disparities by race/ethnicity and socioeconomic status [7–11], as well as past neighborhood-level analyses of pediatric asthma: Harris et al. and Corburn et al. found that asthma hospitalization hotspots in St. Louis and New York City had greater proportions of non-White residents and greater rates of poverty, unemployment, high-density housing, and lack of access to a household vehicle, although they did not test for statistical significance [50,51]. Grunwell et al. found a statistically significant difference between hotspot and non-hotspot census tracts in Georgia for poverty, unemployment, and high-density housing, but not for race [49].

Nearly all patient characteristics in our cohort, including health insurance type and ADI, differed significantly between Black and White patients (Table 4). Most notably, 8.5% of White patients had Medicaid health insurance compared to 39% of Black patients, and the median ADI for White patients was 2.60 (IQR 1.50-4.30) compared to 8.40 for Black patients (IQR 6.50-9.40), making it difficult to assess confounding in our spatial models. Although the VIF indicated that all variables could be included in a multivariable model without substantially inflating variance, collinearity between race and ADI as well as race and health insurance type (S4 Fig) may help explain why ADI and health insurance type were statistically significant in bivariable spatial models but not in the EHR & SEDH-adjusted spatial GAM nor in the EHR & SEDH-adjusted logistic regression (Tables 2 and 3). We attempted to overcome some of these limitations by stratifying our analysis by race. We found that, unlike in the full cohort, adjusting for ADI and health insurance type for Black patients did not attenuate the variance in spatial ORs, suggesting that in our cohort the association between these variables and asthma exacerbations was confounded by race. Our results are consistent with previous observations that racial disparities in asthma control persist even after accounting socioeconomic status [52], but that it is difficult to separate out the effects of socioeconomic status from the effects of race [53]. Confounding of the asthma-socioeconomic status relationship by race has also been observed in past neighborhood-level analyses of asthma morbidity. Zárate et al. found that spatial patterning of asthma-related emergency department visits in Central Texas was partially explained by socioeconomic characteristics in White patients, but not in Black or Hispanic patients [48]. More broadly, understanding the relative contributions of many social determinants of health to asthma is made difficult by their unequal distribution across racial/ethnic groups in the United States [10].

Our health insurance type variable serves as a proxy for individual-level socioeconomic status, and the relationship we measured between it and asthma exacerbation risk is potentially mediated by several pathways including increased rates of smoking [54], psychosocial stress [55], and obesity [56], or an unmeasured variable that varies with socioeconomic status and is also associated with minoritized racial and ethnicity groups in the United States [57]. On the other hand, low neighborhood-level socioeconomic status, which we measured with ADI, is associated with differential exposure to indoor and outdoor air pollution, psychosocial stress from neighborhood violence, and community norms surrounding health behaviors, all of which have been linked to asthma exacerbations [27]. Due to geographic segregation by race that resulted from structural racism and is common in many US cities, including in Philadelphia as we observed in our study, race and neighborhood-level SES are also highly correlated [58]. Thus, our inability to identify an association between asthma exacerbations and ADI when restricting our analysis to Black patients may be due to a restriction of the range of people across the ADI spectrum relative to the range observed in all patients [48]. Future work is needed to understand what specific SEDH variables that covary with race, ADI, and health insurance type are the primary drivers of local disparities in asthma exacerbation risk across Philadelphia.

Our results demonstrate that integration of diverse SEDH datasets with the EHR and the use of both spatial and non-spatial modeling approaches are helpful to understand factors contributing to complex health conditions in real world populations. In both our logistic regression models and spatial GAMs, model fit was not improved by adjusting for SEDH variables in addition to EHR-derived variables. However, in our spatial models, adjusting for SEDH variables resulted in twice as much reduction of the initial variance in spatial ORs compared to adjusting for EHR variables only. Our non-spatial and spatial models also identified different sets of factors associated with exacerbations. For example, ICS prescription was found to have a strong positive association with exacerbations in logistic regression models but did not contribute to the spatial distribution of ORs; conversely, ADI attenuated the variance in spatial ORs more strongly than all other variables tested but was not significant in either logistic regression or negative binomial models. These findings present a framework for future efforts to expand the scope of EHR data, which, especially as the spatial resolution of SEDH datasets continues to increase, will allow for improved individualized exposure estimates. In the future, integration of SEDH data into the EHR may be helpful to tailor asthma management strategies and for health systems to create population-level interventions to improve health of their patients.

This study is strengthened by the high spatial resolution of both our SEDH data and our analysis. Integrating the highest resolution SEDH data available during the time of the study period allowed us to most closely approximate individual-level exposures, and analyzing EHR data at a fine resolution allowed us to understand local health patterns that may not be visible at the census tract or zip-code level. Additional strengths included accounting for many variables and increasing the likelihood that Penn Medicine was patients’ primary care provider by applying SRR restriction. Our study is also subject to limitations, including some related to use of EHR data, such as missingness, entry error, and phenotype misclassification. Additionally, the geocoded addresses used in our study reflect residence at the time of the data pull, but they do not account for residential mobility during the study period, nor does residence information provide a full assessment of environmental exposures.

## Conclusion

By integrating seven datasets containing information on SEDH with an EHR dataset to create individualized exposure assessments, we identified non-spatial and spatial factors associated with asthma exacerbations. Race and prescription of an ICS were most strongly associated with exacerbations in individual-level models. Race also accounted for the most spatial variation in exacerbation odds, along with ADI and health insurance type. Because these three variables had similar spatial distributions, understanding which contributes most to disparities in asthma exacerbations requires additional study of people living in the region identified as a hotspot. Our findings demonstrate how integrating diverse data types and geospatial modeling approaches with EHR data are helpful to understand complex diseases locally.

## Data Availability

The social and environmental determinants of health datasets that support the findings of this study are publicly available in Sensor-based Analysis of Pollution in the Philadelphia Region with Information on Neighborhoods and the Environment (SAPPHIRINE), offered as a web application (http://sapphirine.org) and R package (https://github.com/HimesGroup/sapphirine). Based on ethical and legal considerations, such as that the data was not collected with informed consent, the electronic health record (EHR) data used in this study cannot be shared widely. To request access to EHR data at the Penn Medicine hospital system, reach out to the PennDnA office (https://www.med.upenn.edu/penndna/). The EHR data cleaning methodology is provided as supplementary files to support reproducibility despite being unable to share the data itself.

## Acknowledgements

We would like to thank Sunil Thomas from the University of Pennsylvania Penn Data Store for extracting the EHR data used for this project.

## Supporting information

**S1 Text. Supplementary Methods.**

**S1 Figure. Flowchart of patient cohort selection.** Overview of steps followed to select final patient cohort (N = 6,656) from EHR data on all Penn Medicine patients with at least one asthma ICD code (N = 86,787).

**S2 Figure. Selection of study region using the spatial representation ratio (SRR).** (A) SRR values, defined as the cohort population residing in a block group divided by the underlying population as reported by the 2019 American Community Survey, for all of Philadelphia and for the selected study region (inset box). SRR = 1 indicates no representativeness bias. Density plots of the study cohort (B) before and (C) after filtering for the study region. Base maps were created using the Stamen Design from Stadia Maps.

**S3 Figure. Spatial distribution of SEDH datasets that were integrated with EHR data.** The following maps are shown for the spatial area which comprised our study region: (A) raster of NO2 pollution levels, (B) raster of PM2.5 pollution levels, (C) point sites of toxic releases and the total summed emissions at each site, (D) line segments of roadways and the daily vehicle miles traveled (DVDT) on each, (E) housing violations per block group, normalized by the underlying 2019 American Community Survey population, (F) raster of the normalized difference vegetation index (NDVI). A map of area deprivation index (ADI), which most strongly reduced the spatial variance of odds of exacerbation risk in our spatial GAMs, is shown in Figure 3A. Base maps were created using the Stamen Design from Stadia Maps.

**S4 Figure. Pairwise correlation between all EHR and SEDH variables.** Measures in each box correspond to Pearson’s correlation coefficients. For nominal categorical variables, reference levels are as follows: White (race), Private (health insurance type).

**S5 Figure. Influence of individual EHR and SEDH variables on the odds ratio (OR) of the unadjusted spatial GAM.** Percent reduction in the variance of ORs across the study region for one-variable-at-a-time adjusted spatial GAMs compared to the unadjusted model. OR changes for models adjusted one variable at a time (in addition to years followed) are shown in blue. Multivariable models (both EHR-adjusted and EHR & SEDH-adjusted) are shown in red for comparison.

**S6 Figure. Spatial GAMs adjusted one-at-a-time for the EHR-derived variables that did not greatly reduce variance.** Spatial odds ratios (ORs) of exacerbation are shown after adjusting for years followed and one-at-a-time for the following variables whose percent reduction in variance of ORs was less than 25: (A) age, (B) sex, (C) ethnicity, (D) BMI, (E) smoking status, (F) COPD, (G) allergic rhinitis, (H) Elixhauser comorbidity score, (I) ICS. Base maps were created using the Stamen Design from Stadia Maps.

**S7 Figure. Spatial GAMs adjusted one-at-a-time for the SEDH variables that did not greatly reduce variance.** Spatial odds ratios (ORs) of exacerbation are shown after adjusting for years followed and one-at-a-time for the following variables whose reduction in variance of ORs was less than 25: (A) NO2, (B) PM2.5, (C) toxic releases exposure, (D) vehicular traffic, (E) housing violations, (F) normalized difference vegetation index (NDVI). Base maps were created using the Stamen Design from Stadia Maps.

**S8 Figure. Spatial odds ratios (ORs) of exacerbations among White patients along with the effects of ADI and health insurance type on this distribution.** (A) SRR values for the updated study region used in spatial GAMs for patients of White race only (SRR = 1 indicates no representativeness bias). (B) Unadjusted spatial GAM adjusted only for years followed for patients of White race (N = 1,383). Spatial GAMs adjusted additionally for (C) area deprivation index (ADI) and (D) health insurance type. Base maps were created using the Stamen Design from Stadia Maps.

**S1 Table**. **Generic medication names included in medication classes.** The following generic drug names recorded in the EHR during the study period were used for asthma and exacerbation phenotyping as well as used as independent variables in select models (i.e., ICS). Instances in which these drugs were listed as investigational or nasal formulations were not included.

**S2 Table. Sources and spatiotemporal dimensions of geospatial datasets merged with EHR data.**

**S3 Table**. **Asthma-related housing code violations extracted from the Philadelphia Department of Licenses and Inspections.**

**S4 Table**. **Patient medications by exacerbation count levels.** Shown are the number and percentage of patients receiving each of the medication types listed according to their number of exacerbations during the study period.

**S5 Table. Characteristics of complete cases and patients excluded due to missingness.** Shown are the number and percentage of patients in each level for categorical variables and the Median and Interquartile Range (IQR) for the Years followed variable in complete cases versus those excluded due to missingness in the sex, ethnicity, health insurance type, BMI, and smoking status variables.

**S6 Table. Adjusted Generalized Variance Inflation Factors (GVIFs) for each EHR and SEDH variable included in the EHR & SEDH-adjusted negative binomial and logistic regression models.**

**S7 Table**. **Individual-level asthma exacerbation risk factors in multivariable negative binomial regression models.** Shown are the adjusted incidence rate ratios (IRRs), 95% confidence intervals (CIs), and p-values for negative binomial models of asthma exacerbations as a count outcome adjusted for EHR-derived variables only and for both EHR-derived and SEDH variables.

**S8 Table. Spatial GAMs of asthma exacerbations adjusted for individual risk factors that most changed risk**. Shown are the adjusted odds ratios (ORs), 95% confidence intervals (CIs), and p-values for spatial GAMs of asthma exacerbations as a dichotomous outcome adjusted for years followed and one-at-a-time for ADI, race, and health insurance type, the three variables whose percent reduction in variance of ORs was greater than 25.

